# Rare Germline Variants and Genetic Susceptibility to Pleural Mesothelioma: Moving Beyond DNA Repair

**DOI:** 10.64898/2026.09.29.26364318

**Authors:** Elton Jalis Herman, Carla Debernardi, Khadija Sana Hafeez, Sara Devito, Tze Yin Lim, Alessia Russo, Alessandra Allione, Elisabete Weiderpass, Antonio Agudo, Marta La Vecchia, Irma Dianzani, Natalia Cabrera Castro, Vittorio Simeon, Rosario Tumino, Marcela Guevara, Laia Peruchet-Noray, Ana Jiménez-Zabala, Paolo Vineis, Elisabetta Casalone, Giuseppe Matullo

**Affiliations:** Department of Medical Sciences, University of Turin, Via Santena 19, 10126 Turin, Italy; Division of Nephrology, Department of Medicine, Columbia University, New York, NY, USA; International Agency for Research on Cancer, World Health Organization, 69372 Lyon, France; Unit of Nutrition and Cancer, Catalan Institute of Oncology-ICO, 08908 L’Hospitalet de Llobregat, Spain; Interdepartmental Center for Studies on Asbestos and Other Toxic Particulates "G. Scansetti", University of Turin, 10126 Turin, Italy; Department of Health Sciences, University of Eastern Piedmont, Novara, 28100, Italy; Department of Epidemiology, Murcia Regional Health Council, 30007 Murcia, Spain; Research Group on Epidemiology and Public Health, Biomedical Research Institute of Murcia Pascual Parrilla (IMIB), 30120 Murcia, Spain; Consortium for Biomedical Research in Epidemiology and Public Health (CIBERESP), 28029 Madrid, Spain; Medical Statistics Unit, University of Naples "L. Vanvitelli", Naples, 80138, Italy; Hyblean Association for Epidemiology Research, A.I.R.Ep. ETS Ragusa, Italy; Instituto de Salud Pública y Laboral de Navarra, 31003 Pamplona, Spain; Centro de Investigación Biomédica en Red de Epidemiología y Salud Pública (CIBERESP), 28029 Madrid, Spain; Navarra Institute for Health Research (IdiSNA), 31008 Pamplona, Spain; Cancer Epidemiology and Prevention Research Unit, Department of Epidemiology and Biostatistics, School of Public Health, Imperial College London, Wood Lane, London W12 0HZ, United Kingdom; Biogipuzkoa Health Research Institute, Group of Epidemiology of Chronic and Communicable Diseases, San Sebastian, Gipuzkoa, Spain; Ministry of Health of the Basque Government, Sub-Directorate for Public Health and Addictions of Gipuzkoa, San Sebastian, Gipuzkoa, Spain; MRC Centre for Environment and Health, Imperial College London, St Mary’s Campus, London, UK; Medical Genetics Unit, AOU Città della Salute e della Scienza, 10126 Turin, Italy

**Keywords:** Pleural Mesothelioma, Whole Exome Sequencing, Rare Germline Variants, European Prospective Cohort, Precision Medicine, Polygenic Background

## Abstract

A minority of asbestos-exposed individuals develop Pleural mesothelioma (PM), suggesting a role for inherited genetic factors. We performed germline whole-exome sequencing of blood from 127 EPIC participants who developed PM and controls. Rare pathogenic/likely pathogenic variants were assessed in 193 cancer-associated genes and were evaluated for association with PM risk and overall survival. Exome-wide gene-based rare-variant aggregation analyses were performed using SKAT-O and Firth logistic regression. A PM polygenic risk score (PRS) was integrated with PTV status and asbestos exposure. PTVs were enriched in cases and associated with increased PM risk (OR=3.5, 95% CI 1.3–11.2, p-value = 0.01). PTV carriers had improved survival and lower mortality after adjustment for age at diagnosis and sex (HR=0.54, 95% CI 0.32–0.92, p-value = 0.023). PTV carriers had lower PRS values, and among individuals with low asbestos exposure, they were associated with increased PM risk (OR=5.75, 95% CI 1.09–58.2, p-value = 0.023). Gene-based analyses identified four novel loci. Rare variants and polygenic background may jointly influence PM risk, however validation in larger cohorts is warranted.

## Introduction

The importance of environmental carcinogens in cancer development has been well established through extensive epidemiological and mechanistic studies [1]. Individuals environmentally exposed to asbestos, or workers involved in asbestos product manufacturing, face an increased risk of developing asbestos-related pulmonary diseases and severe malignancies such as the rare pleural mesothelioma (PM) later in life [2]. Today, PM remains an aggressive and therapeutically refractory malignancy with dismal prognosis, characterized by a median overall survival of 12–18 months despite current treatment modalities [3–5]. While chronic asbestos exposure represents the primary etiological factor, the disease exhibits substantial interindividual susceptibility disparities that cannot be attributed to exposure alone. Remarkably, PM has a characteristically low mutational burden [6] and not all individuals who undergo the exposure subsequently manifest an asbestos-related disease [40].

Germline pathogenic variants in *BAP1* cause BAP1-tumor predisposition syndrome and increase the risk of mesothelioma and other malignancies [7]. They occur in approximately 1–2% of unselected mesothelioma cases, while higher frequencies in selected familial series likely reflect ascertainment bias [6, 9, 10]. More broadly, pathogenic variants in *BAP1* and other cancer-predisposition genes, including *ATM*, *BARD1*, *BRCA2*, *BRIP1*, *XPC*, and *CHEK2* are found in approximately 10–12% of unselected patients, supporting a wider heritable component of PM susceptibility and highlighting the importance of genetic counselling and surveillance [9–12, 14].

In the updated ASCO Guidelines for PM treatment, genetic counselling has been recommended for all individuals with a diagnosis of PM [13]. Germline mutations in *BAP1* and DNA repair genes demonstrate significantly improved survival and possibly impacting therapeutic responses [15–17]. Current studies are often limited by small sample sizes or focused gene panels, which may fail to capture the full heterogeneity of rare and underrepresented pathogenic variants.

Beyond high-penetrance variants in cancer predisposition genes, common genetic variation also contributes to cancer risk through a polygenic component that can be summarised using polygenic risk scores (PRSs). We previously investigated the utility of PRS for PM risk prediction in a large cohort of PM retrospective cases and reported that PRS showed greater predictive performance among individuals with low asbestos exposure [31]. Complementary work has demonstrated that individuals whose observed phenotype deviates from their genetically expected value based on PRS are enriched for rare, deleterious variants in genes linked to early-onset forms of disease, supporting the idea that rare and common variants can exert counterbalancing or synergistic effects on liability [25]. However, the relative contributions of rare high-impact variants, common polygenic burden, and environmental exposure to PM risk remains unexplored.

To address these gaps, our study leverages Whole Exome Sequencing (WES) to analyse pre-clinical PM (n = 127) cases in the EPIC cohort providing an unbiased assessment of germline coding variants and offering new insights into the heritable components that confer high or moderate risk for this rare malignancy. We also investigated whether combining potentially pathogenic variants across genes, can offer a more comprehensive picture of individual susceptibility. This approach may yield important biological insights into tumour-causation mechanisms, emphasising the importance of the germline genomic background in cancer predisposition.

## Results

### Population features of study subjects

The primary analysis included 127 individuals who subsequently developed PM, each matched to a non-cancer control by age, sex, asbestos exposure grade, and histotype. The characteristics of the study population are summarised in Table EV1. Sequencing achieved a mean depth of 75.0 ± 4×, with an average of 82 ± 7% of target regions covered at >50× (Table EV3, Appendix Figure S1). A schematic representation of the analysis workflow is depicted in Fig. 1.

**Figure 1.**
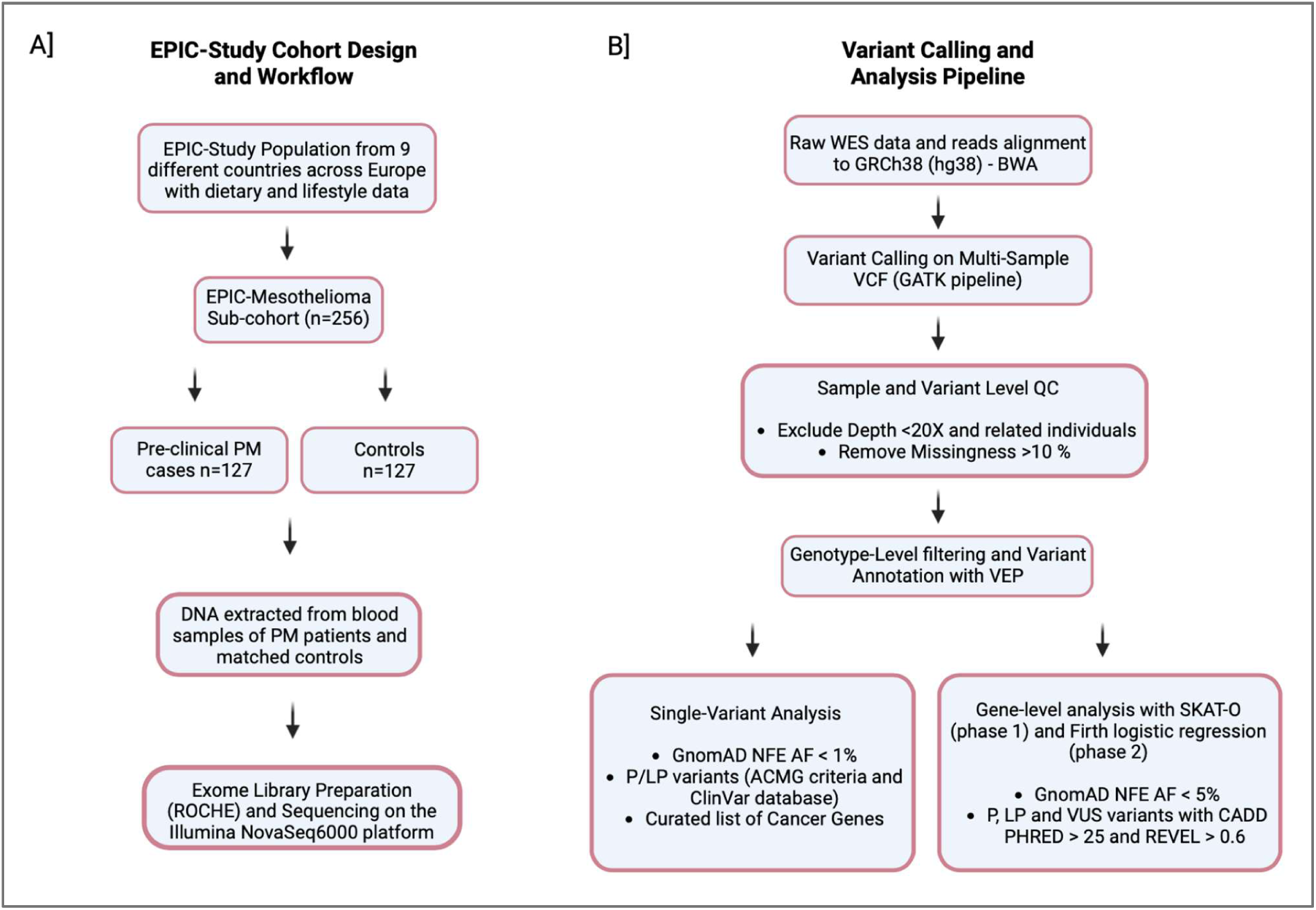
- **EPIC-Mesothelioma Study Cohort Design [A] and Analysis Pipeline [B].** From the main EPIC cohort, participants who developed pleural mesothelioma during follow-up in the EPIC cohort were individually matched to cancer-free controls to generate the EPIC-Mesothelioma sub-cohort. Germline DNA from pre-diagnostic blood samples underwent whole-exome sequencing, followed by alignment, variant calling, quality control, and annotation. The workflow then diverged into two analytical branches: single-variant analysis of rare pathogenic and likely pathogenic variants in a curated literature-based cancer-gene panel, and gene-based aggregation analysis of rare deleterious variants across the exome. The latter included a discovery analysis in EPIC-Mesothelioma and a confirmation analysis using EPICOR non-cancer controls. **Abbreviations:** PM, pleural mesothelioma; WES, whole-exome sequencing; GATK, Genome Analysis Toolkit; VCF, Variant Call Format; BWA, Burrows-Wheeler Aligner; VEP, Variant Effect Predictor; QC, quality control; AF, allele frequency; P/LP, pathogenic/likely pathogenic; ACMG, American College of Medical Genetics and Genomics; VUS, variant of uncertain significance; CADD, Combined Annotation Dependent Depletion; SKAT-O, Sequence Kernel Association Test-Optimal.

### Pre-clinical Pleural Mesothelioma Cases with Pathogenic and Likely Pathogenic Germline Variants

As a first step, we focused on a manually curated literature-based panel of 193 cancer-associated genes (Table EV2) to assess the burden of pathogenic and likely pathogenic (P/LP) germline variants in pre-clinical PM cases and controls. Among PM cases, we identified 29 P/LP variants spanning 27 genes (Fig. 2; Table EV4). Variant classes were predominantly protein-altering and included 12 missense mutations (41.4%), 7 stop-gained mutations (24.1%), 6 frameshift mutations (20.7%), 3 splice-site mutations (10.3%), and 1 start-loss mutation (3.4%).

**Figure 2.**
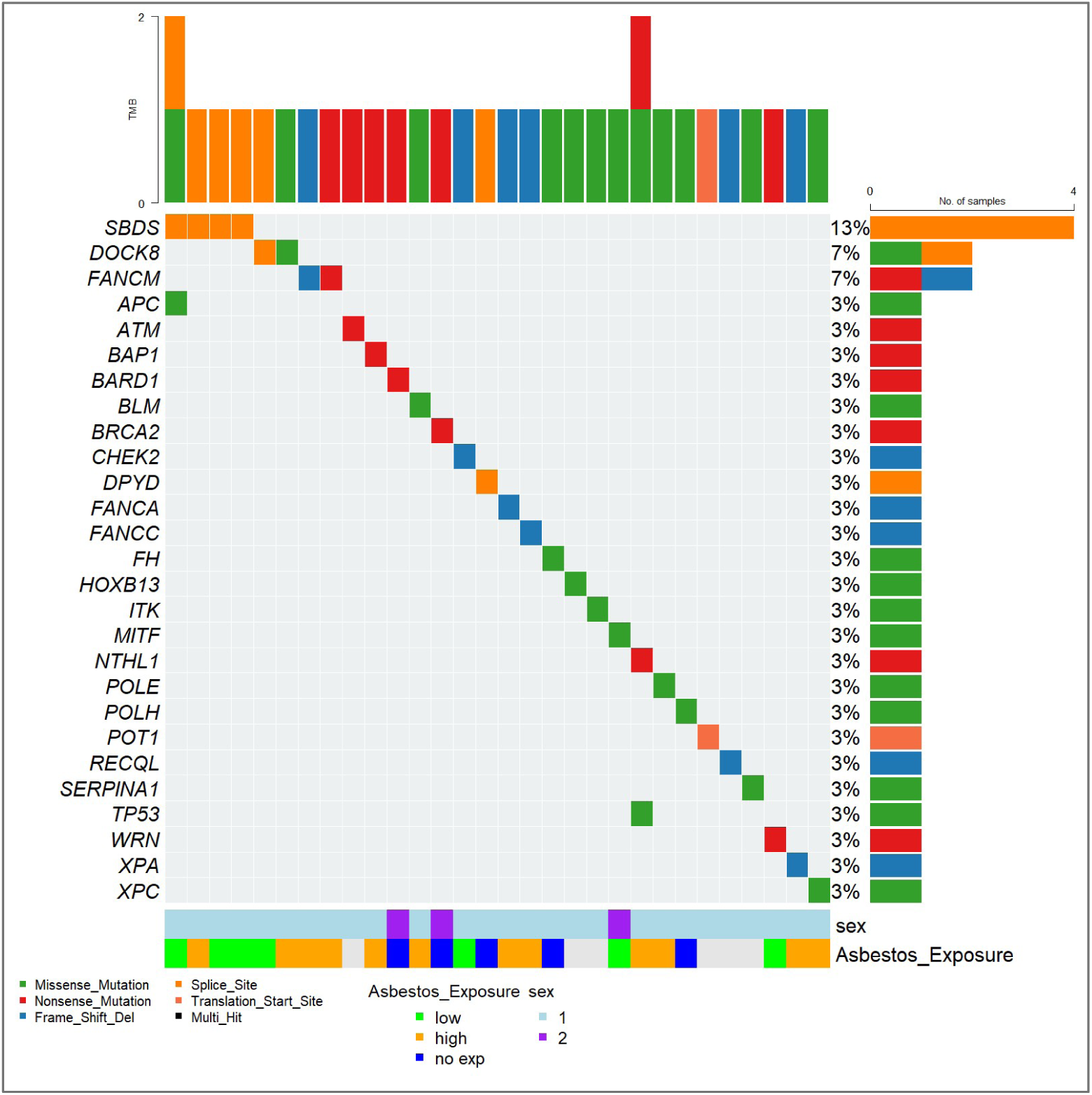
**Molecular landscape of pathogenic and likely pathogenic (P/LP) germline variants in pre-clinical pleural mesothelioma cases**. Oncoplot displaying the distribution and characterization of P/LP germline variants across 30 pre-clinical pleural mesothelioma patients. Each column represents an individual patient, and the height of stacked bars indicates the total number of variants per patient (TMB: tumor mutation burden). Variant classes are colour-coded as indicated. Individual gene mutation frequencies displayed as rows, with each row representing a distinct gene. Bottom annotation tracks display patient demographics - asbestos exposure status (categorical: high, low, no exposure) and biological sex (1: male, 2: female).

We observed a higher burden of rare pathogenic or predicted deleterious variants in PM cases compared with controls (Fisher’s exact test, p = < 0.001; OR = 4.6, 95% CI: 1.9-12.1). Overall, 31 PM patients (24%) carried P/LP variants in cancer-associated genes, representing a significantly higher proportion compared with controls (6%). *ATM*, *BRCA2*, *FANCM*, *HOXB13*, *MITF*, *RECQL*, *SBDS* and *DOCK8* were recurrently mutated across the entire cohort of patients and controls with *FANCM*, *SBDS*, and *DOCK8* each mutated in more than one case (Table EV4).

When restricting the analysis to protein-truncating variants (PTVs), we identified such variants in 20 pre-clinical PM patients across 16 genes (Table EV4). Carriers of PTVs in cancer predisposition genes showed an approximately more than three-fold increased risk of PM compared with controls (Fisher’s exact test, p = 0.01; OR = 3.5; 95% CI: 1.3–11.2).

Specifically, 24% patients carried P/LP variants compared with 6% of controls, whereas non-carriers accounted for 76% and 94% of these groups, respectively. These findings indicate a significant enrichment of PTV carriers among PM cases, supporting an association between germline truncating variation in cancer predisposition genes and increased PM risk (Appendix Figure S2). Notably, PTVs were detected in genes associated with tumor predisposition syndrome: a stop-gain mutation in *BAP1* was found in an individual with high asbestos exposure and a start-loss variant in *POT1* was reported in a PM patient without a documented occupational asbestos exposure. Additional mutations are described in Table EV4.

### Survival Analysis

We evaluated the association between PTVs in cancer-associated genes and overall survival in patients with pleural mesothelioma. In the primary analysis, in which non-carriers included individuals carrying missense variants, PTV carriers showed longer overall survival than non-carriers (log-rank p-value = 0.048) (Fig. 3A). Multivariable Cox regression adjusted for age at diagnosis and sex showed that PTV carriers had a lower mortality risk (HR = 0.54, 95% CI: 0.32–0.92, p-value = 0.023).Across approximately 54 months of follow-up, the survival curves remained separated, with higher estimated overall survival among PTV carriers. At 24 months after diagnosis, estimated overall survival was approximately 50% (95% CI: 32.3–77.5%) among PTV carriers compared with 22.9% (95% CI: 16.0–32.6%) among non-carriers.

**Figure 3.**
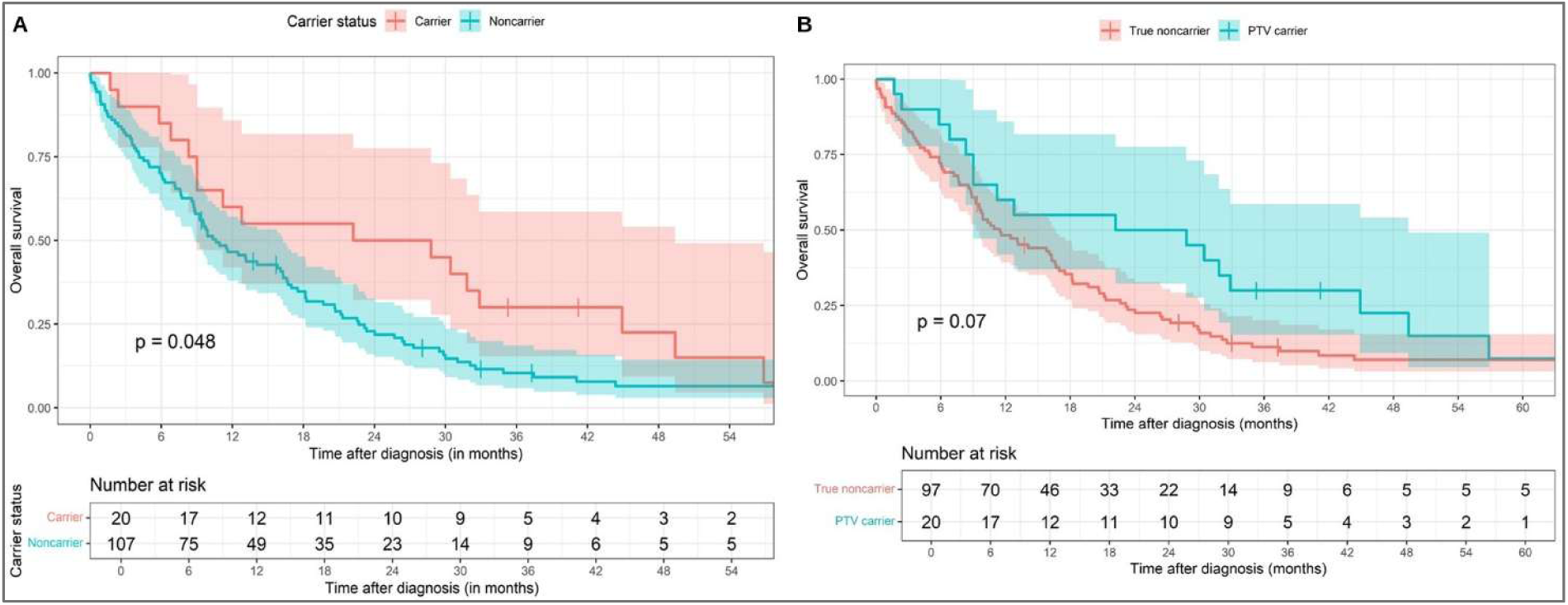
Kaplan–Meier overall survival according to germline protein-truncating variant (PTV) carrier status in patients with pleural mesothelioma. **(A)** Kaplan–Meier analysis comparing overall survival between PTV carriers and non-carriers, with the non-carrier group including individuals with missense variants. The analysis includes follow-up from diagnosis, with numbers at risk shown at 0, 6, 12, 18, 24, 30, 36, 42, 48 and 54 months. **(B)** Kaplan–Meier analysis comparing overall survival between PTV carriers and true non-carriers, excluding individuals with missense variants. Numbers at risk are shown at 0, 6, 12, 18, 24, 30, 36, 42, 48, 54 and 60 months.

To further evaluate whether this association was driven by inclusion of missense variant carriers in the non-carrier group, we performed the analysis restricted to PTV carriers versus true non-carriers, excluding individuals with missense variants. In this analysis, PTV carriers continued to show higher estimated overall survival, although the difference between the Kaplan–Meier curves did not reach conventional statistical significance by the log-rank test (p-value = 0.07) (Fig 3B). In contrast, multivariable Cox regression adjusted for age at diagnosis and sex showed a lower mortality risk among PTV carriers (HR = 0.57, 95% CI: 0.33–0.97, p-value = 0.0367). An analysis of missense variants alone versus true non-carriers did not show a significant association with overall survival (Figure EV1). Together, these analyses support an association between germline PTV carrier status and overall survival that is observed both when missense variant carriers are included in the comparison group and when they are excluded.

### Polygenic Risk Score and Rare Variant integration

Building on our previously evaluated PRS in the EPIC-MESO cohort [30], we next leveraged this framework to test whether integrating PRS with PTVs could improve PM risk stratification. We observed that PM cases carrying PTVs had significantly lower PRS than non-carrier cases (p = 0.04), suggesting that rare high-impact variants may contribute to disease susceptibility even in the presence of a lower polygenic burden. Further, stratifying by asbestos exposure, PTV carriers in the no/low exposure group had significantly increased PM risk compared with non-carriers (OR 5.75, 95% CI 1.09–58.2, p = 0.023), although the wide confidence interval reflects the small number of carrier controls (Table EV10) and within this low-exposure group, PM cases carrying rare PTVs (N = 6) showed lower PRS than non-carrier cases (N = 18; p = 0.04), whereas no differences were observed in the no- and high-exposure groups (Figure EV2).

## Gene-Collapsing Analysis reveals Additional Susceptibility Loci

Beyond the effect of individual variants on disease risk, the aggregation of rare and low frequency variants can act jointly to further increase susceptibility to the disease. We performed a gene collapsing analysis in EPIC-MESO dataset to investigate potential genes harbouring at least two variants in genes that may increase the cancer risk. We selected rare and low-frequency variants classified as pathogenic and likely pathogenic according to ACMG and Clinvar classification and predicted pathogenic variants (CADD > 20, REVEL > 0.6) as described in Material and Methods. We tested 2,823 genes carrying 4,011 qualifying variants adjusting for sex, age and the first two principal components (PCs). SKAT-O analysis, at a nominal significance threshold (p < 0.05), identified 141 gene loci (Fig. 4, Table EV5).

**Figure 4.**
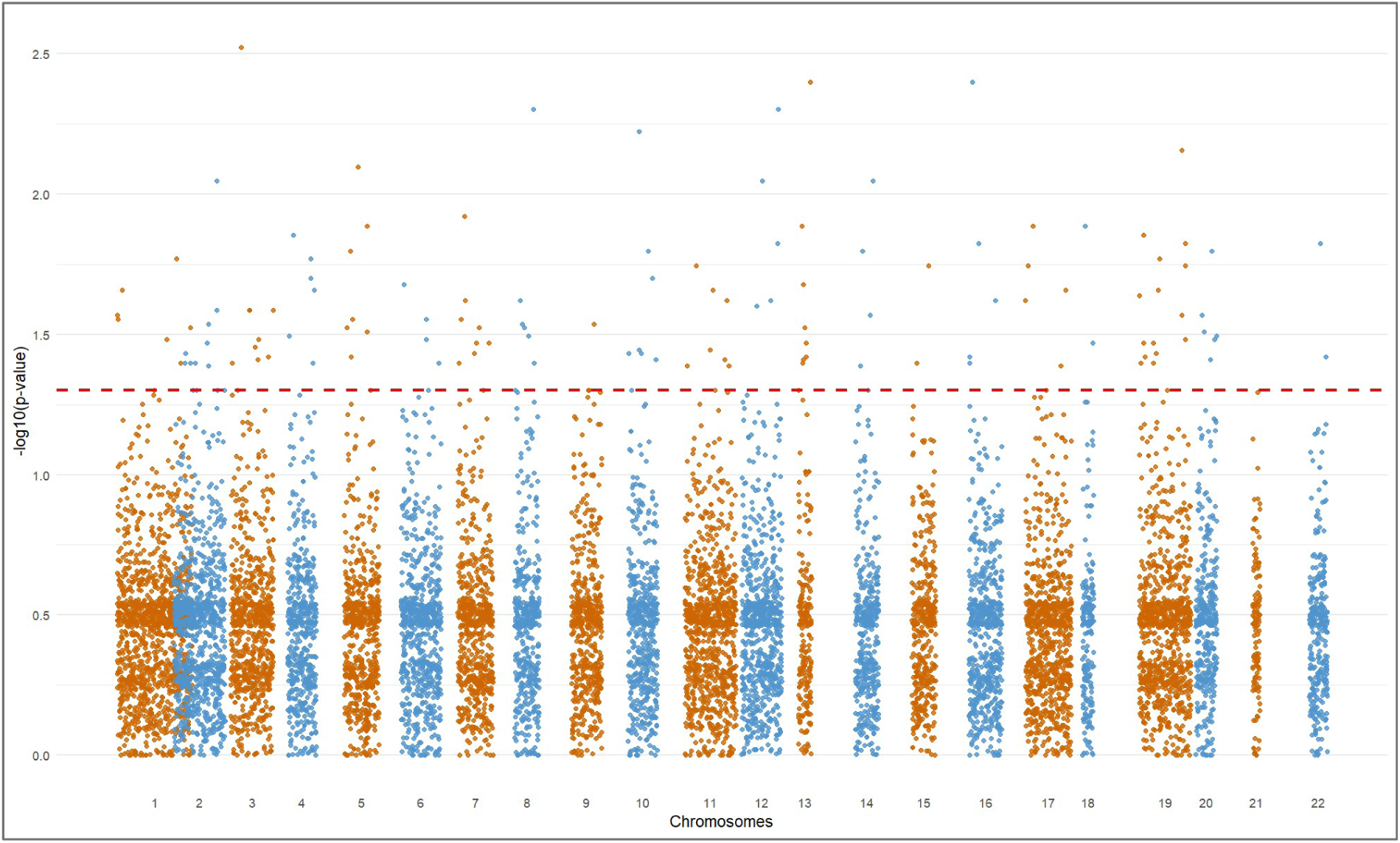
- **Gene-based Collapsing Analysis in the EPIC-MESO cohort.** Manhattan plot showing exome-wide results from the SKAT-O analysis of rare and low-frequency variants for pleural mesothelioma risk. Each point represents a gene, plotted according to chromosomal position on the x-axis and −log10(SKAT-O P-value) on the y-axis. The dashed red line corresponds to a nominal P-value threshold of 0.05.

Then, we assessed the cumulative effect of qualifying variants (QV) on PM risk by aggregating variants at the gene level in EPIC-MESO cases and comparing them with an independent set of non-cancer individuals from another prospective cohort [28]. Joint variant calling followed by quality control and filtering retained 3,162 QVs across 1,355 genes for the analysis. Gene-level burden was evaluated using Firth logistic regression, restricted to genes with at least two carriers and adjusted for sex, age, and the first two PCs.

Although no gene remained significant after false discovery rate (FDR) correction, 36 genes showed nominally significant enrichment of QVs in PM cases compared to non-cancer controls (Table EV6). Among these, *UNC13D*, *SHANK2*, *TLCD2*, and *GMPR2* exhibited the highest frequency of rare and low-frequency QV carriers in PM cases (Figure 5). Notably, we confirmed significant enrichment for *SUCLA2* (p = 0.01, OR = 18.4), *UNC13D* (p = 0.02, OR = 18.4, OR = 5.6), *TCF7L2* (p = 0.03, OR = 29.9), and *COL28A1* (p = 0.0005, OR = 204.8), consistent with findings from the initial analysis using the European cohort as controls (Figure 5 and 6; Table EV6). When applying a null model restricted to synonymous variants, no genes were significantly enriched, confirming that the observed signals in the primary analysis are unlikely to be driven by inflation or technical artifacts (Table EV7).Gene enrichment analysis of 36 genes with the highest frequency of QV carriers in PM revealed a significant overrepresentation of metabolic pathways involved in nucleotide biosynthesis and redox processes (Table EV8), particularly GO:0003920 (GMP reductase activity) and GO:0046857 (oxidoreductase activity acting on other nitrogenous compounds with NAD or NADP as acceptors).

**Figure 5.**
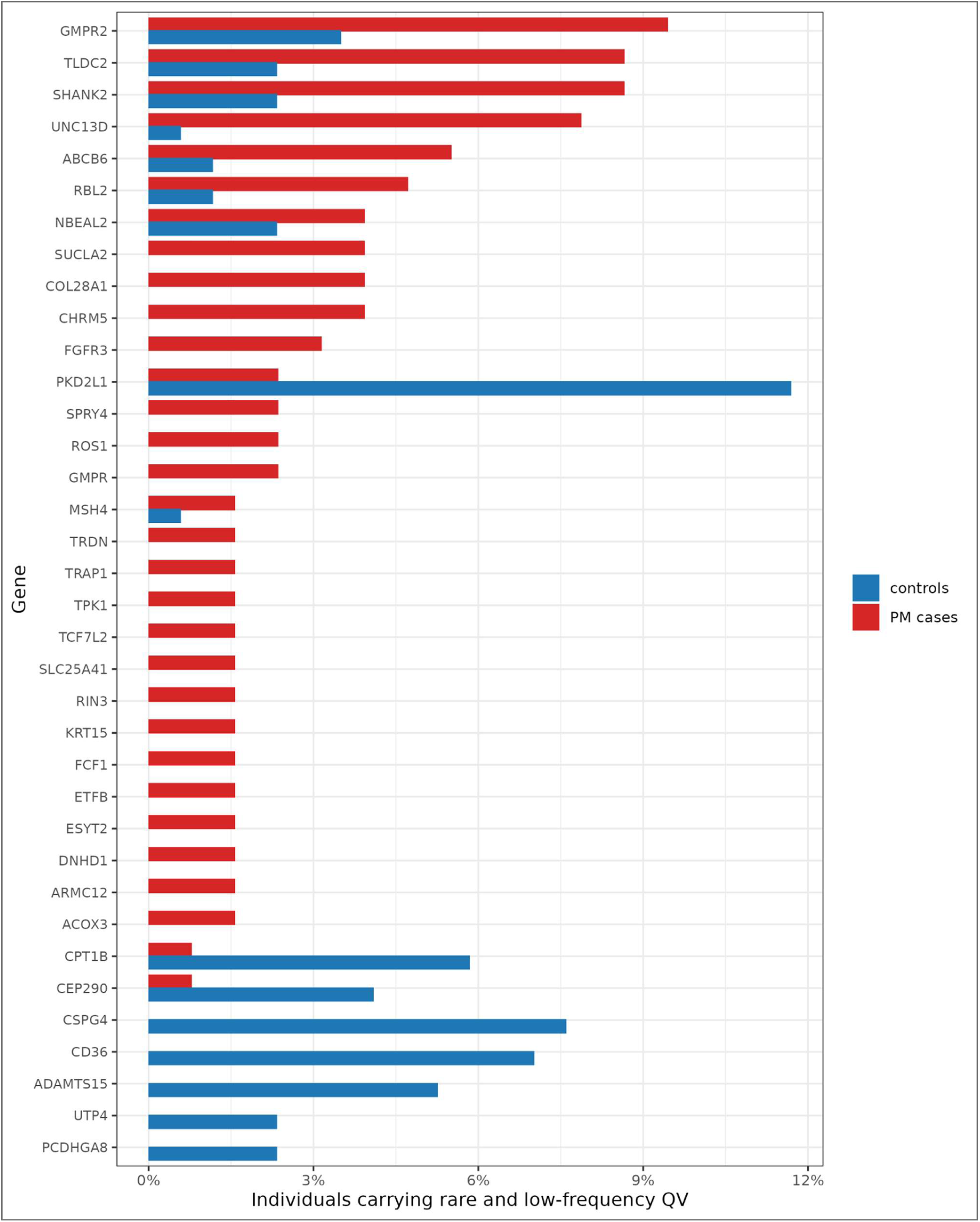
Frequency of carriers of rare and low-frequency qualifying variants in PM cases and EPICOR non-cancer controls. Bar plot showing the percentage of individuals carrying rare and low-frequency qualifying variants in each gene among PM cases and EPICOR non-cancer controls. Red bars represent PM cases and blue bars represent EPICOR controls. Genes shown are those included in the gene-based burden analysis with qualifying variants observed in either group.

**Figure 6.**
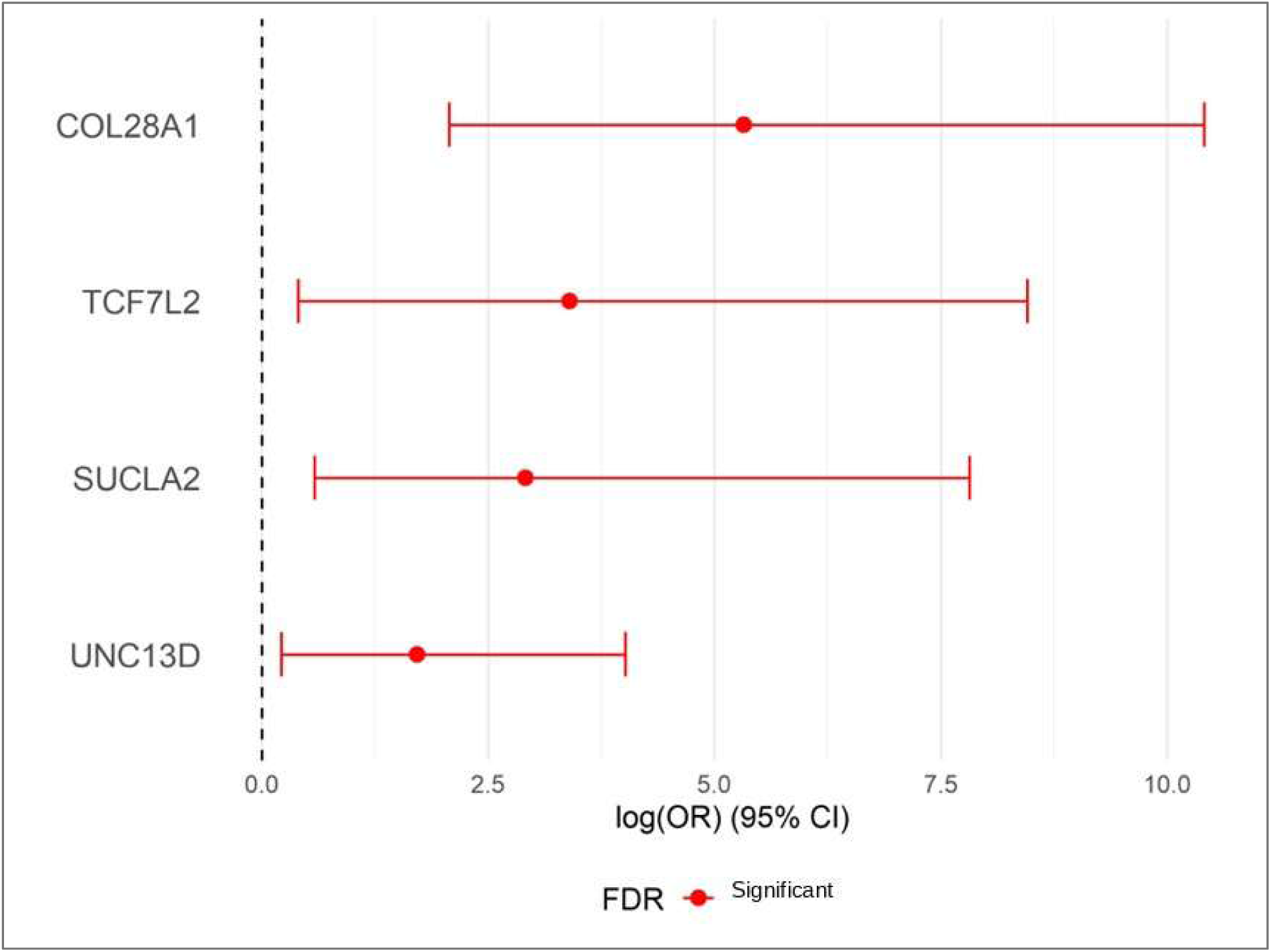
Gene-based burden confirmation analysis in EPIC-MESO cases and EPICOR controls. Forest plot showing odds ratios and 95% confidence intervals for the association between aggregated rare and low-frequency qualifying variants and pleural mesothelioma risk in the four genes identified by the SKAT-O analysis. Effect estimates were obtained using Firth’s penalized logistic regression and are displayed on the log(OR) scale. FDR-significant associations are indicated.

## Discussion

Our WES study investigates the contribution of rare germline pathogenic variants to PM risk and supports the view that patients with PM are more genetically heterogeneous than previously reported. To address this, we employed two complementary analytical approaches: the first aimed at identifying highly penetrant, clinically impactful mutations, and the second, focused on detecting rare and low-frequency risk variants that, when aggregated at the gene level, may contribute to disease susceptibility through non-canonical biological pathways. By leveraging an exome-wide approach in a European cohort, our study expands the current understanding of germline genetic genetic architecture of PM, extending beyond prior studies capturing common variation, or targeted NGS gene panels with limited genomic scope [39, 45].

Single-variant analysis identified rare P/LP variants in 27 cancer-associated genes, accounting for 24% of PM cases and conferring a significantly increased risk compared to non-cancer controls (OR = 4.6, p = 0.0002). These findings are consistent with previous evidence highlighting the involvement of key DNA damage response and repair pathways, including homologous recombination (*ATM*, *BAP1*, *BARD1*, *BLM, BRCA2*), nucleotide excision repair (*POLE*, *XPA*, *XPC*), cell cycle checkpoint regulation (*CHEK2*, *TP53*), and the Fanconi anemia pathway (*FANCA*, *FANCC*, *FANCM*), underscoring the central role of genomic maintenance mechanisms in PM susceptibility.

Although in this prospective study the analysis of PM patients carrying PTVs or P/LP variants did not reveal associations across the different asbestos exposure categories, asbestos remains the primary etiological factor for PM. The lack of significant associations may reflect the challenges of accurately assessing asbestos exposure based on questionnaire-derived information, which do not capture the cumulative dose of exposure.

Nevertheless, these findings are consistent with the hypothesis that germline defects affecting DNA repair, genome maintenance, telomere integrity, immune regulation, or metabolic pathways may influence individual susceptibility to asbestos-induced carcinogenesis. Asbestos exposure promotes persistent oxidative stress, DNA damage, and chromosomal instability, and inherited defects in these biological processes could impair the cellular response to such insults, thereby facilitating the progressive accumulation of genomic alterations that may contribute to mesothelial transformation. In this context, the identification of pathogenic or likely pathogenic variants in genes such as *BAP1* and *POT1* is of particular interest, as both genes play key roles in maintaining genome integrity and represent biologically plausible candidates for modulating interindividual asbestos risk to PM. We identified a pathogenic *BAP1* variant (p.Tyr401Ter) in exon 12, which has not previously been reported in PM but has been described as a somatic alteration in uveal melanoma [40]. PM harboring *BAP1* mutations are often characterized by reduced aggressiveness and increased sensitivity to chemotherapy, potentially due to compromised DNA repair capacity and consequent reliance on alternative repair mechanisms [19–21]. Consistent with this observation, the carrier of this variant was diagnosed at 54 years of age and exhibited a survival exceeding three years, supporting prior evidence linking germline *BAP1* mutations with improved prognosis in PM [19–21]. Although the relatively early onset may suggest familial *BAP1* tumor predisposition syndrome, unfortunately detailed clinical information was not available for this individual within the EPIC cohort.

Similarly, we identified a germline *POT1* variant, another gene involved in genome maintenance through telomere protection. Germline *POT1* variants have been associated with several hereditary cancer predisposition syndromes (POT1–TPD). The encoded protein is a key structural component of the shelterin complex as it shields the single-stranded telomeric overhang preventing aberrant activation of the DNA damage response at chromosome ends [42]. Heterozygous somatic *POT1* mutations were initially identified in chronic lymphocytic leukaemia cells, and germline mutations were later reported in families with isolated cancers, including melanoma and glioma [43]. Mutations in *POT1* have been identified in multiple cancer types and are estimated to have an overall prevalence of up to 3%. The *POT1* c.1A>G variant disrupts the translation initiation codon of the POT1 mRNA and is predicted to interfere with *POT1* protein synthesis and in particular this variant has been reported in the published literature in an individual affected with multiple myeloma [44]. This observation further supports the hypothesis that rare germline variants, particularly when considered collectively at the gene or pathway level, may contribute to PM development through biological mechanisms extending beyond canonical DNA repair pathways. We identified the truncating *BRCA2* variant p.Lys944Ter in a female participant with no known occupational asbestos exposure who subsequently developed biphasic PM. This stop-gained variant has been reported in individuals and families with breast, ovarian, and pancreatic cancer, supporting its established clinical relevance in BRCA2-associated hereditary cancer predisposition syndromes [45–48]. Germline pathogenic variants in *BRCA2* have also been reported in mesothelioma cohorts and may contribute to PM susceptibility, although their prevalence in unselected mesothelioma populations appears low and remains imprecisely estimated. Detailed information on the participant’s personal and family history of cancer, as well as tumor-based evidence of a second hit or homologous-recombination deficiency, was unavailable; therefore, the contribution of this variant to PM development in this individual cannot be determined.

Genes involved in telomere protection and genome stability, including *POT1, WRN* and *RECQL* contribute to the preservation of chromosomal integrity. Their dysfunction may lead to telomere attrition, replicative stress, and chromosomal rearrangements, processes frequently observed in PM. Recent works have suggested that germline *BLM* mutations significantly increase susceptibility to PM in asbestos-exposed individuals, likely by promoting genomic instability, impairing apoptosis in mesothelial cells and enhancing proinflammatory responses in macrophages [41]. Consistent with this evidence, we identified a missense variant in the *BLM* functional domain in a male patient with high asbestos exposure and epithelioid PM.

These findings are consistent with accumulating evidence suggesting that germline defects in DNA repair and damage response pathways, including genes involved in telomere protection and genome stability, may contribute to cancer development in the context of asbestos exposure [45]. Beyond these well-characterized pathways, we identified additional P/LP variants in lesser-known genes spanning immunological functions (*DOCK8, ITK*), metabolic pathways (*DPYD, FH, SERPINA1*), transcriptional control (*HOXB13, MITF*), WNT signaling (*APC*), *SBDS* (ribosomal function) and specialized DNA repair mechanisms (*POLH, NTHL1, WRN, RECQL*), thereby expanding the genetic architecture of PM susceptibility.

Additional genes, including *FH*, *MITF*, *HOXB13*, and *SERPINA1*, point to the contribution of metabolic dysregulation, transcriptional control, and chronic inflammatory processes in modulating mesothelioma risk. For instance, *FH* mutations may promote tumorigenesis through metabolic reprogramming and oxidative stress, potentially synergizing with asbestos-induced cellular damage [50].

Current studies on integrating both rare and common variants improves the identification of high-risk individuals [25]. In our stratified analyses, PTV carriers in the low-PRS group showed an increased estimate of PM risk within low occupational asbestos-exposed people, while no difference was observed in the other asbestos groups, highlighting the role of the fiber in PM onset. These data support a model in which rare germline variants and polygenic burden may jointly modulate PM risk, with genetic effects appearing more pronounced at lower asbestos exposure, consistent with gene– environment interaction frameworks described in cancers [55]. However, given the small numbers and wide, overlapping confidence intervals, these patterns should be regarded as hypothesis-generating and require confirmation in larger, independent cohorts.

Another important aspect of this study is the association between PTVs and improved survival. This finding suggests that germline background may influence not only PM risk but also disease trajectory after diagnosis. One possible explanation could be that tumors arising in carriers of PTVs may be more vulnerable to DNA damage and therefore show enhanced sensitivity to first-line chemotherapy. Another possibility is that these PM tumors follow a distinct evolutionary trajectory leading to less aggressive clinical profile. The survival benefit should be interpreted with caution due to the reduced number of individuals at risk at later time points, but it raises the possibility that PTV germline variant status could eventually serve as a prognostic stratification indicator to help define biologically meaningful groups for systemic treatment planning by stratifying patients more likely to benefit from chemotherapy or immunotherapy. These findings may have relevant clinical implications and could help refine PM diagnosis through the development of a hereditary PM gene panel.

Future analyses should not be restricted to established cancer-predisposition genes, but should also be extended to additional genes emerging from genome-wide or exome-wide aggregation approaches. This broader strategy may capture genes involved in biological processes not traditionally classified as cancer-related, including immune regulation, mitochondrial metabolism, extracellular matrix remodeling, and tumor–microenvironment interactions. Expanding the analysis beyond canonical cancer genes may therefore improve the identification of inherited factors that influence PM susceptibility, tumor evolution, treatment response, and clinical outcome in a larger cohort.

Rare and low frequency variants within the same gene, although genetically distinct, may exert similar functional consequences by disrupting the same protein function or molecular pathway. Aggregating these variants at the gene level enables the assessment of their cumulative contribution to disease susceptibility and improves the detection of gene-based associations. We performed gene-collapsing analysis to determine the cumulative effect of pathogenic and potentially pathogenic variants on PM risk. The gene-based approach led to the identification of *SUCLA2*, *TCF7L2*, *COL28A1*, and *UNC13D* associated with PM in two different cohorts of European and Italians non-cancer individuals. In line with previous gene-level rare-variant aggregation studies, these genes may highlight biological pathways potentially contributing to cancer susceptibility. Even if *TCF7L2* has never been described de-regulated in PM, it maps to the canonical Wnt signaling pathway, implicated in mesothelioma cell proliferation and survival. Aberrant activation of Wnt/B-catening signaling promotes tumor aggressiveness and therapeutic resistance [51]. In contrast, *SUCLA2* is not established as a hereditary cancer predisposition gene but has emerging relevance in tumor metabolism, as it encodes a mitochondrial enzyme involved in the tricarboxylic acid cycle; alterations in mitochondrial metabolism are increasingly recognized as contributing to tumor growth and therapeutic resistance [52]. *UNC13D* is primarily associated with immune regulation, and its deficiency impairs cytotoxic lymphocyte function, potentially increasing susceptibility to malignancies particularly lymphomas through compromised immune surveillance [53]. Finally, evidence linking *COL28A1* to cancer remains limited, although its role as a collagen family member suggests a possible contribution to extracellular matrix remodeling and tumor cell behavior [54]. Overall, these findings suggest a heterogeneous set of mechanisms, spanning canonical oncogenic signaling, metabolic adaptation, immune dysfunction, that may collectively contribute to cancer risk or progression.

Both moderate and high-risk germline variants have been associated with an increased risk of disease; however, current evidence indicates that these variants are not sufficient to induce PM in the absence of asbestos or other carcinogenic mineral fiber exposure. Rather, they are likely to modulate the individual response to asbestos-induced carcinogenesis within a gene–environment interaction framework. The unavailability of tumor tissue, which prevented us from assessing whether the identified germline variants were associated with loss of protein expression or other functional consequences in the corresponding tumors. Moreover, the lack of clinical informationare important limitations of this study. Nevertheless, the reliability of the findings is supported by the robustness of the analytical model, which accounted for potential confounders and genomic inflation.

## Conclusion

Our findings further support this model by identifying rare germline variants affecting genes involved in DNA repair, genome maintenance, and telomere integrity, highlighting biological pathways that may contribute to PM development in the context of asbestos exposure or other carcinogenic insults. Although larger studies are required to clarify the contribution of individual variants, comprehensive germline profiling may improve our understanding of PM pathogenesis and help identify individuals who could benefit from tailored surveillance strategies. In addition, the identification of inherited alterations may have therapeutic relevance by informing the development of personalized treatment approaches, including strategies exploiting tumor-specific synthetic lethality.

## Materials and Methods

### Study cohort

The European Prospective Investigation into Cancer and Nutrition (EPIC) cohort comprises over 520,000 individuals who were recruited between 1992 and 2000 from 23 study centers across 9 European countries (Denmark, France, Germany, Italy, Norway, Spain, Sweden, the Netherlands, and the UK). The study design has been described previously [26]. The EPIC study is a large-scale, detail-oriented, long-term cohort study that aims to investigate the relationship between diet, lifestyle, and environmental factors and the incidence of cancer and other chronic diseases. From the main EPIC cohort, participants who developed pleural mesothelioma (PM) during 20 years of follow-up were identified and individually matched for sex, age, center of recruitment and asbestos exposure with cancer-free controls. As previously described, in the EPIC-Mesothelioma (EPIC-MESO) sub-cohort 135 participants developed PM after a mean follow-up of 8.3 years (range 0.5-18.8 years) [27]. The disease was classified by ICD-10 diagnosis codes (ICD, International Statistical Classification of Diseases, Injuries and Causes of Death) as C45.0 (Mesothelioma of Pleura) in all participants. Cancer endpoint data is based on the latest round of follow-up received from the EPIC centres and centralised at IARC between 2014 and 2016 [29–31]. Asbestos exposure was assessed from occupational information available in the baseline EPIC questionnaire [29, 31]. A semiquantitative job-exposure matrix (JEM) was developed by an expert epidemiologist to assess the probability and intensity (0 = no exposure, 1 = low exposure, 2 = high exposure) of exposure to asbestos [27, 28].

## Cancer free controls

For the validation of gene-level analysis findings, the EPICOR nested case–control study was used as an independent control cohort to assess the generalizability and performance of the genetic association models [32]. The genetic study included 584 Italian participants (data from 171 samples were used in this study) who developed coronary artery disease (CAD) during follow-up, along with matched control subjects.

## Whole exome sequencing (WES) and data analysis

Genomic DNA was extracted from whole blood samples as previously described [27, 28]. WES was performed on available DNA samples derived from 128 PM pre-clinical cases, 128 non-cancer controls nested within the EPIC MESO study and from 171 EPICOR samples. The Library Preparation was done using the KAPA HyperExome kit (Roche, Basel, Switzerland) with 150 ng of input DNA per sample. Libraries sequencing was performed on an Illumina NovaSeq 6000 sequencer (paired-end 2×150 bp read configuration).

The obtained sequenced reads were evaluated with Fastqc (v.0.11.9) and aligned to the human genome assembly GRCh38 by employing the Burrows-Wheeler Aligner software (0.7.18). Single-nucleotide variants (SNVs), small insertions and deletions (indels) were called following Genome Analysis Toolkit (v.4.1.9.0) Best Practices workflow using HaplotypeCaller. For EPIC-MESO and EPICOR per-sample genomic variant call format (gVCF) files were generated and jointly genotyped using GATK GenotypeGVCFs to produce a multi-sample VCF file. Splitting of multiallelic variants was performed on joint gVCF (bcftools v.1.21), followed standard hard filtering according to GATK Best Practices recommendations. The variants were called using canonical MANE select transcripts and annotated using Ensembl Variant Effect Predictor (VEP) to assess functional impact (frameshift, stop-gain, stop-loss, missense, in-frame indel, and splice site variants) and predicted deleteriousness using in silico prediction tools (PolyPhen-2, SIFT, CADD, REVEL, ClinPred, DANN, MetaLR, PrimateAI scores). The potential impact of variants on splicing was predicted using SpliceAI (https://spliceailookup.broadinstitute.org/). Clinical classifications of variants were obtained from the ClinVar database (https://www.ncbi.nlm.nih.gov/clinvar). Population allele frequencies were annotated using data from the Genome Aggregation Database (gnomAD, v4.1.0). The pathogenicity of the identified variants in each patient was assessed according to the American College of Medical Genetics and Genomics and the Association for Molecular Pathology (ACMG/AMP) guidelines [34] using GeneBe software [35].

## Sample and variant quality controls

In EPIC-MESO four individuals were excluded based on standard sample-level metrics, including excess relatedness (identity-by-descent [IBD] > 0.125) and heterozygosity, in accordance with criteria previously applied in the EPIC-MESO study [27]. Samples were required to have a genotyping call rate greater than 90%. Variants with a missingness rate exceeding 10% were excluded. To ensure high-confidence genotype calls for downstream analyses, variants were subjected to stringent quality control and filtering based on variant allele frequency (VAF), read depth (DP), and mapping quality metrics. Genotypes were classified according to VAF thresholds in conjunction with sequencing depth as follows: genotypes with VAF between 0.30 and 0.70 and DP ≥ 20 were classified as heterozygous; and genotypes with VAF ≥ 0.90 and DP ≥ 20 were classified as homozygous alternate. Genotypes with VAF < 30 inconsistent with expected allele balance were classified as uncertain and those genotypes supported by fewer than 20 reads (DP < 20), irrespective of VAF, were set to missing and excluded from downstream analyses to minimize genotype misclassification. This conservative filtering strategy was adopted to reduce false-positive calls and ensure robustness of variant-level and gene-level analyses. Following quality controls and filtering, a total of 595,878 high-confidence variants across 20,832 genes were retained for subsequent analyses for 254 individuals.

### Selection of qualifying variants for single variant analysis and gene-level Association Analysis

To focus on rare variants with a potential higher impact on PM, only variants with < 1% allele Frequency in the gnomAD Non-Finnish European (gnomAD NFE) population were retained for subsequent analysis and variants classified as benign or likely benign by ClinVar were excluded. Furthermore, intron, and synonymous variants along with certain variants with varying functional impacts were also excluded from further analysis (Table EV9).

QVs were then selected based on the pathogenic criteria indicated by ACMG/AMP guidelines implemented in GeneBe (https://docs.genebe.net/pl/), obtaining 1,730 Pathogenic (P) and Likely Pathogenic (LP) rare variants from canonical MANE select transcripts in 1,371 genes.

As an initial step, single rare variant analysis was performed by extracting QV within a manually curated literature-based list of 193 genes selected as cancer hallmark genes associated with hereditary cancers and implicated in PM (Table EV2). Precedence was given to variants affecting the protein-coding sequence (Frameshift, Splice Acceptor, Splice Donor, Start Lost and Stop Gained mutations) and according to the ClinVar database and ACMG/AMP guidelines. The P and LP variants of interest were manually inspected in integrated genome browsers (IGV) and BAM files to confirm accurate genotype calling and exclude potential alignment artefacts.

For downstream gene-based analysis, we selected rare and low-frequency variants (gnomAD exome NFE AF < 0.05) predicted to be P or LP, supplemented by *in silico* prediction scores. Specifically, variants were retained if they met at least one of the following criteria: (i) ACMG P/LP; (ii) ClinVar LP with CADD PHRED > 25; (iii) ClinVar Conflicting classifications of pathogenicity (including “association” or “risk factor”) with CADD PHRED > 25; (iv) ClinVar Pathogenic/Likely Pathogenic and risk factor with CADD PHRED > 25; or (v) no ClinVar annotation with CADD PHRED > 25 and REVEL > 0.6.

The analysis was conducted in two phases with independent datasets. Phase 1 (n = 254 samples) involved the discovery EPIC cohort comparing pre-clinical PM cases with matched controls. Phase 2 (n = 298 samples) consisted of validation analyses utilizing an independent control cohort derived from the EPICOR study which comprised individuals with documented cardiovascular outcomes, including myocardial infarction, and coronary revascularization. The use of cardiovascular disease-affected controls in the validation phase provides a stringent negative control population, enabling assessment of whether identified gene-based associations are specific to PM susceptibility.

In phase 1, gene-based association testing was performed using the Sequence Kernel Association Test-Optimal (SKAT-O), implemented in the R SKAT package [36–38].

For each gene, SKAT-O combined the association contributions of all QV (loss-of-function and missense) with the binary disease outcome (case versus control status), while adjusting for age, sex, and the first two PCs. P-values were calculated by fitting null models containing only covariates, enabling efficient genome-wide computation.

In phase 2, collapsing analysis was performed using Firth’s bias-reduced logistic regression, as implemented in the logistf R package (https://CRAN.R-project.org/package=logistf), with EPIC-MESO cases and EPICOR as controls. Analysis was restricted to genes with at least two variant carriers. For each gene, carrier status (presence vs absence of QV) was tested for association with PM susceptibility. To assess potential sources of test statistic inflation, we built a null model restricted to rare synonymous variants. The same collapsing framework and regression model were applied, testing gene-level carrier status based on synonymous variants. The distribution of association statistics from this analysis was then evaluated to verify calibration and to detect any systematic inflation.

Statistical significance was assessed using likelihood ratio tests derived from the Firth regression framework. Multiple testing correction was applied using the FDR method. Confidence intervals and P-values were computed with the profile penalized log likelihood method. Genes of interest were plotted as a forest plot.

## Sanger Sequencing

The mutation *SBDS* c. 258+2 T>C identified by WES in four cases was validated through Sanger sequencing. Briefly, the genomic region of interest was amplified by PCR using AmpliTaq Gold™ 360 Master Mix (Thermo Fisher Scientific, USA) with primers designed to flank the variant site and to specifically amplify the *SBDS* gene rather than its pseudogene (SBDSex2intF 5’ AGTGTTTGTAAATGTTTCTA 3’ and SBDSex2bisR 5’ CtggaggaaagactaaccaagC 3’). PCR products were purified using ExoSAP-IT™ PCR Product Cleanup Reagent (Thermo Fisher Scientific, USA) and subjected to cycle sequencing using fluorescently labeled dideoxynucleotides using the BigDye Terminator v3.1 Cycle Sequencing Kit (Applied Biosystems, USA), according to the manufacturer’s instructions. Sequencing reactions were run on an ABI 3500 Genetic Analyzer (Applied Biosystems, USA).

## Statistical and survival Analysis

The proportions of individuals with one or more P/LP QV in the cancer genes list, were compared with those from the control group with Fisher’s Exact test. Overall survival was defined as the time from diagnosis to death or censoring. Follow-up time was calculated from the date of diagnosis to the earliest of date of death, last follow-up (2016), or administrative censoring (December 31, 2017). Individuals without a recorded death before the censoring date were treated as censored. Kaplan-Meier survival curves were generated to compare survival distributions between groups, and differences were assessed using the log-rank test. Multivariable Cox proportional hazards regression models were fitted to estimate hazard ratios (HRs) and 95% confidence intervals (CIs), adjusting for age at diagnosis and sex. The proportional hazards assumption was evaluated using Schoenfeld residuals.

## Integration of polygenic risk and rare protein-truncating variants

To assess whether common polygenic burden and rare PTV carriage represent independent or overlapping contributions to PM risk, we integrated an internal Polygenic risk score (PRS) with PTV carrier status in the EPIC-Meso exome subset. PTV carrier status was tested against case/control status using Fisher’s exact test within asbestos-exposure strata (no/low exposure] vs high exposure). All statistical analysis was done in R statistical software (4.2.1). Further details are available in the method details for the specific analysis.

## Data Availability

The data supporting the findings of this study are available from the corresponding author upon reasonable request, following approval by the EPIC Data Access Committee. Access to the EPIC data is governed by the EPIC access policy, as detailed in https://epic.iarc.fr/access/

## Ethics Statement

The study was approved by the Institutional Review Board of IARC and the ethics committees in the participating countries (reference number: IEC 24-08). All participants gave their written informed consent to participate in the EPIC study. The present study was carried out in accordance with the ethical principles of the Declaration of Helsinki.

## CRediT authorship contribution statement

Elton Jalis Herman: Conceptualization; methodology; data curation; formal analysis; visualization; validation; writing – original draft; project administration; resources.

Carla Debernardi: Validation; writing – review and editing; data curation.

Tze Yin Lim: Data curation; writing – review and editing; formal analysis.

Khadija Sana-Hafeez: Data curation; writing – review and editing; formal analysis.

Sara Devito: Data curation; writing – review and editing; formal analysis.

Alessia Russo: Methodology; validation; data curation; writing – review and editing

Alessandra Allione: Data curation; writing – review and editing.

Elisabete Weiderpass: Writing – review and editing; data curation; resources

Antonio Agudo: Writing – review and editing; data curation; resources

Marta La Vecchia: Data curation; resources; writing – review and editing

Irma Dianzani: resources; writing – review and editing.

Natalia Cabrera Castro: Data curation; resources; writing – review and editing.

Vittorio Simeon: Data curation; resources; writing – review and editing.

Rosario Tumino: Data curation; resources; writing – review and editing.

Marcela Guevara: Data curation; resources; writing – review and editing.

Laia Peruchet-Noray: Data curation; resources; writing – review and editing.

Ana Jiménez-Zabala: Data curation; resources; writing – review and editing.

Paolo Vineis: Data curation; resources; writing – review and editing; conceptualization.

Elisabetta Casalone: Writing – review and editing; software; data curation; supervision; conceptualization; project administration.

Giuseppe Matullo: Conceptualization; supervision; project administration; writing – review and editing; funding acquisition.

## Acknowledgements

The coordination of EPIC is financially supported by the International Agency for Research on Cancer (IARC) and also by the Department of Epidemiology and Biostatistics, School of Public Health, Imperial College London which has additional infrastructure support provided by the NIHR Imperial Biomedical Research Centre (BRC). The authors acknowledge the National Institute for Public Health and the Environment (RIVM), Bilthoven, Netherlands, for their contributions and ongoing support to the EPIC Study. The authors also acknowledge the contribution of Dario Mirabelli, Department of Medical Sciences, Unit of Cancer Epidemiology, CPO Piemonte and University of Turin, 10126 Turin, Italy for his invaluable contribution in the assessment of asbestos exposure. The national cohorts are supported by: Danish Cancer Society (Denmark); Ligue Contre le Cancer, Institut Gustave Roussy, Mutuelle Générale de l’Education Nationale, Institut National de la Santé et de la Recherche Médicale (INSERM) (France); German Cancer Aid, German Cancer Research Center (DKFZ), German Institute of Human Nutrition Potsdam-Rehbruecke (DIfE), Federal Ministry of Education and Research (BMBF) (Germany); Associazione Italiana per la Ricerca sul Cancro-AIRC-Italy, Compagnia di SanPaolo and National Research Council (Italy); Dutch Ministry of Public Health, Welfare and Sports (VWS), Netherlands Cancer Registry (NKR), LK Research Funds, Dutch Prevention Funds, Dutch ZON (Zorg Onderzoek Nederland), World Cancer Research Fund (WCRF), Statistics Netherlands (The Netherlands); Health Research Fund (FIS)—Instituto de Salud Carlos III (ISCIII), Regional Governments of Andalucía, Asturias, Basque Country, Murcia and Navarra, and the Catalan Institute of Oncology—ICO (Spain); Swedish Cancer Society, Swedish Research Council and County Councils of Skåne and Västerbotten (Sweden); Cancer Research UK and the Medical Research Council (United Kingdom).

## Expanded View Figure Legends

**Figure EV1. Kaplan–Meier overall survival according to missense-only germline variant carrier status.** Kaplan–Meier analysis comparing overall survival between true non-carriers and individuals carrying missense variants only. Overall survival was followed from diagnosis, with 95% confidence intervals shown around the survival estimates. The number at risk is indicated below the survival plot. The difference between the groups was not statistically significant by the log-rank test (p-value = 0.12). In multivariable Cox regression adjusted for age at diagnosis and sex, missense-only carriers showed an HR of 1.75 (95% CI: 0.87–3.51; p-value = 0.11) compared with true non-carriers.

**Figure EV2. Polygenic risk score according to PTV carrier status and asbestos exposure. (A)** Distribution of the internal polygenic risk score (PRS) according to PTV carrier status among pleural mesothelioma cases and controls. Boxplots show the internal PRS for carrier and non-carrier groups among cases and controls; sample sizes are indicated below each group. P-values for the within-group comparisons are shown above the corresponding brackets. **(B)** Distribution of the internal PRS according to PTV carrier status stratified by asbestos exposure category (no exposure, low exposure, and high exposure) among PM cases and controls. Boxplots show the internal PRS for PTV carriers and non-carriers within each exposure group; sample sizes are indicated below each group, and P-values for the within-group comparisons are shown above the corresponding brackets.

